# An internet-assisted sleep, dietary, and physical activity intervention to support weight-loss among postpartum people (Sleep GOALS): Protocol for a pilot randomized controlled trial

**DOI:** 10.1101/2025.03.25.25324617

**Authors:** Marquis S. Hawkins, Esa M. Davis, Kaleab Z. Abebe, Kathleen M. McTigue, Namhyun Kim, Mariska Goswami, Daniel J. Buysse, Judy C. Chang, Michele D. Levine

## Abstract

**Background:** Postpartum weight retention and maternal obesity are associated with short- and long-term maternal morbidity and mortality risk. Most weight-loss interventions among postpartum individuals follow evidence-based lifestyle recommendations but have produced only modest effects and had substantial heterogeneity. We developed a novel internet-assisted weight management intervention for postpartum people that integrates concepts for improving sleep health within a diet and physical activity-focused intervention. We describe the intervention protocol and discuss how the pilot study’s findings will inform future development and evaluation.

**Methods:** We will recruit 40 postpartum individuals with overweight or obesity from Western Pennsylvania to participate in a single-blind, parallel-arm, randomized controlled trial design. Participants will be randomized at a 1:1 ratio to the Sleep GOALS (Goal-focused Online Access to Lifestyle Support) intervention or education control group. The Sleep GOALS intervention includes interactive lessons addressing sleep, diet, physical activity, behavioral self-monitoring tools, and a lifestyle coach to provide accountability, encouragement, and personalized support. The education control will receive brochures from the American Academy of Sleep Medicine (e.g., sleep hygiene, sleep in women), SNAP education connection (e.g., family-friendly activities, meal planning), and the U.S. Department of Health and Human Services (e.g., physical activity promotion during and after pregnancy). Primary study outcomes include the intervention feasibility (i.e., recruitment, enrollment, attrition rates, intervention engagement) and acceptability (i.e., participant ratings of the intervention delivery, curricula, approach to behavioral self-monitoring, action plans, intervention platform, and coaching). Secondary outcomes include weight loss and retention of pregnancy and postpartum weight gain.

**Discussion:** Incorporating a holistic approach that addresses sleep health alongside diet and physical activity, the Sleep GOALS intervention aims to not only facilitate weight loss but also enhance overall maternal well-being. Pilot testing will help us identify and refine factors related to the conduct of the planned larger, definitive trial and estimate the change in secondary outcomes.

## Background

Obesity is a global health concern that disproportionately affects birthing people of reproductive age. ^1, 2^ In the US, approximately two-thirds of birthing people of reproductive age have overweight or obesity. ^1, 2^ Postpartum weight retention (PPWR) significantly contributes to obesity development, independent of pre-pregnancy body mass index (BMI) and gestational weight gain. ^3–6^ In the short term, PPWR is associated with an increased risk of maternal complications like gestational diabetes and hypertensive disorders of pregnancy in subsequent pregnancies. Long-term, PPWR is associated with obesity development and increased risk for developing obesity-related cardiovascular disease (e.g., hypertension, diabetes). ^6–8^ Thus, the postpartum period is critical for engaging birthing individuals in weight-loss interventions to prevent PPWR and cardiovascular disease morbidity and mortality.

The US Preventive Service Task Force (USPSTF) recommends clinicians offer patients with obesity weight-management interventions that provide counseling on diet and physical activity. ^9^ However, results of interventions focused only on postpartum diet and physical activity have been modest. ^10–12^ For example, a diet and exercise intervention involving 66 postpartum women revealed that nearly half did not achieve their 5% weight loss objectives. ^13^ Other trials have similarly reported high variability, with many women achieving little to no weight loss. ^14, 15^ Person-related factors common during the postpartum period, such as sleep disturbances, likely contribute to the modest impact of these interventions. ^16–18^

Epidemiological and experimental evidence links sleep to weight, weight gain, and fat metabolism in the general population. Research on the independent influence of sleep and PPWR is sparse but generally shows an inverse relationship between sleep duration and PPWR and maternal obesity. ^19–22^ Poor sleep health can reduce the efficacy of weight-loss interventions through behavioral (e.g., diet and physical activity) ^23–36^ and biological mechanisms (e.g., fat metabolism). ^37^ Yet, diet and physical activity weight management interventions rarely integrate sleep interventions. An exception is the Better Weight-Better Sleep Study, which randomized 49 participants to a diet and exercise weight loss intervention with (BWBS) or without an integrated sleep intervention. ^38^ Participants in the BWBS intervention lost twice as much weight as those in the control group. Although this study provides preliminary evidence that addressing sleep can enhance the effectiveness of diet and physical activity interventions on weight loss, the impact of an integrated sleep and weight management program among postpartum individuals specifically has not been evaluated.

Considering the existing research gaps, we developed Sleep GOALS (Goal-focused Online Access to Lifestyle Support), an internet-assisted sleep, diet, and physical activity weight-loss intervention for postpartum people. To develop Sleep GOALS, we used a patient-centered approach to adapt and integrate content from an existing sleep (Transdiagnostic Sleep and Circadian; TSC) and physical activity and diet weight loss intervention (GOALS). TSC is a modular intervention that addresses sleep health, rather than sleep disorder, making it broadly applicable for public health interventions. GOALS, developed by Dr. McTigue and colleagues, is an online version of the CDC-recognized Diabetes Prevention Program’s Lifestyle Intervention.^39^ This pilot study aims to determine the interventions’ feasibility and acceptability. The secondary aim is to estimate the interventions’ preliminary efficacy on postpartum weight retention and weight loss. Here, we describe the trial protocol and how findings will inform the subsequent treatment development and evaluation phase.

## Methods

### Study design and population

This study uses a single-blind, parallel-arm, pilot randomized control trial (RCT) design. We will include females between 2 and 7 months postpartum with overweight or obesity (i.e., BMI ≥25 kg/m^2^). We chose this postpartum range because postpartum weight retention during this period is associated with long-term obesity development.^5, 6^ Additionally, at three months postpartum, most infants experience increases in nocturnal sleep.^40^ Thus, many of our behavioral strategies for improving maternal sleep may be more feasible at this point rather than earlier in the postpartum period. It is safe for most birthing people to participate in an intervention involving physical activity during this period.^41^ However, we will screen for contraindications for physical activity using a standard questionnaire,^42^ and require approval from a healthcare provider before enrollment if contraindications are identified. We will further restrict our sample to physically inactive people (<150 min/week) who report at least one indicator of poor sleep health (i.e., regularity, satisfaction, alertness, timing, efficiency, and duration) based on the RU SATED questionnaire.^43^ We will enroll all women of reproductive age. We anticipate that most participants will be between 15 and 49 years of age. Table 2 describes the additional inclusion/exclusion criteria.

**Table 1.**
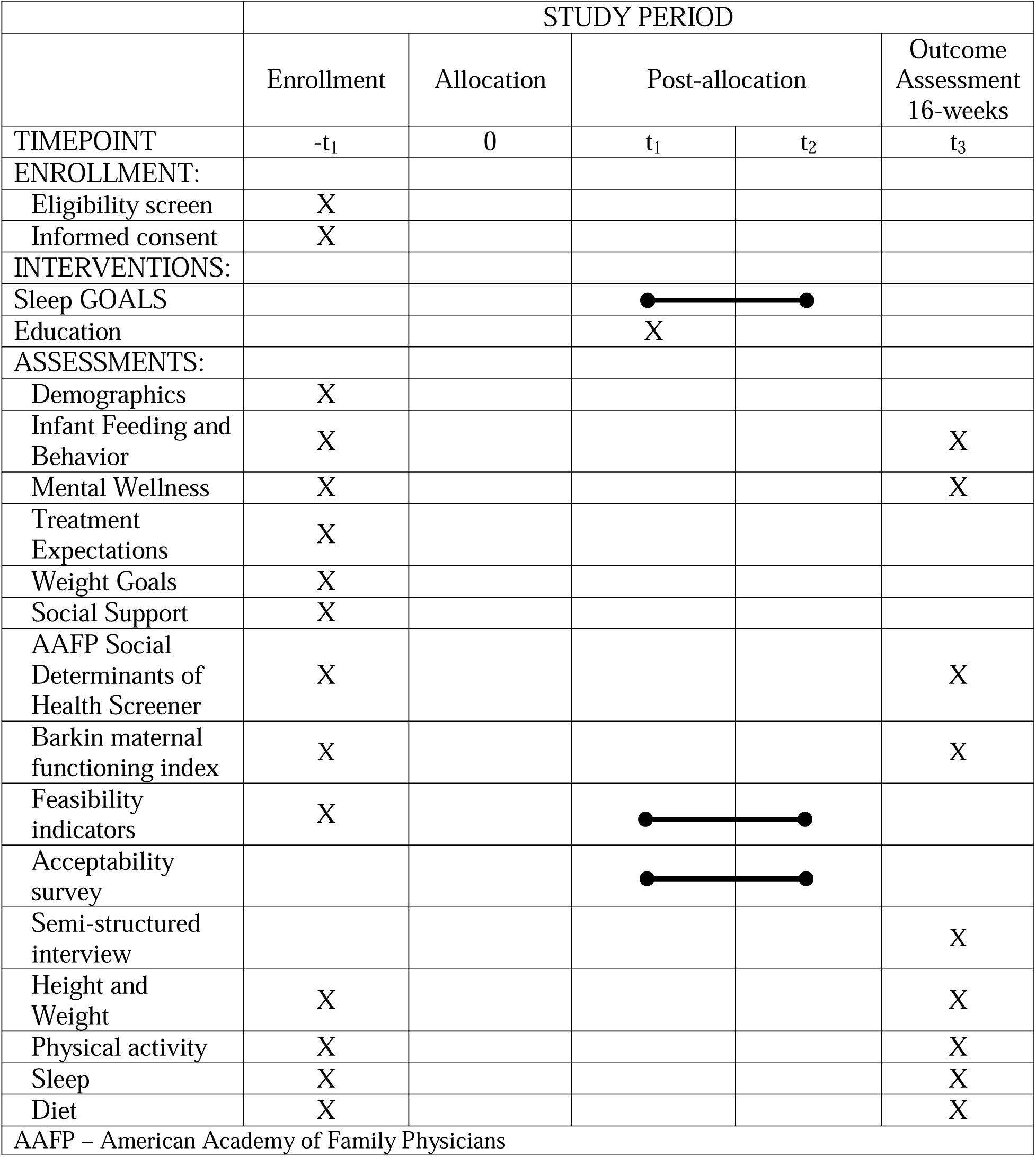
Schedule of enrolment, interventions, and assessments.

**Table 2.** Inclusion/exclusion criteria.

|  |
| --- |
| Table 2 – Inclusion/exclusion criteria |
| Inclusion criteria |
| 1. Primiparous women 2 to 7 months postpartum |
| 2. BMI 25 to 40 kg/m<sup>2</sup> |
| 3. Physically inactive (<150 min/week) |
| 4. Endorse $\geq 1$ indicator of poor sleep health based on the RU_SATED questionnaire |
| 5. Internet access (computer or phone) |
| Exclusion criteria |
| 1. Currently pregnant or planning to become pregnant during the study period |
| 2. Medications that affect weight (e.g., steroids) or sleep |
| 3. Participating in another weight-loss intervention |

### Recruitment and Enrollment

Our recruitment goal is to enroll 40 participants. We will attempt to recruit a diverse sample and not exclude participants based on race or ethnicity. However, we anticipate the sample will be predominantly black/African American and non-Hispanic white based on the demographic characteristics of Allegheny County. The 2019 Census data reports that approximately 80% of Allegheny County residents identify as white, and approximately 13% identify as black/African American. Only 2.2% of Allegheny County residents identify as Hispanic or Latino. We will attempt to oversample (>30%) black/African American women since African Americans are disproportionately burdened by sleep-related obesity. Since Black-identifying individuals are disproportionately affected by maternal obesity and postpartum weight retention, we will attempt to over-sample in this population (≥30%).^44, 45^ All participants will be recruited from Allegheny County, PA, using 1) Pitt+Me, an extensive clinical research registry maintained by the University of Pittsburgh’s Clinical and Translational Science Institute,^46^ 2) University electronic mailings, 3) BuildClinical, an online research recruitment company, 4) Pittsburgh Brown Mama’s participant services, 5) targeted postcard mailings to participant registered in the Magee Obstetric Materna and Infant database and biobank,^47^ and 6) direct clinician-engagement through the Newborn Research Support Services (NuRSERy) program.^48^ Interested participants will be pre-screened for eligibility online or through a registry call center. Referrals to contact potentially eligible subjects will be delivered through the registry’s online portal. A study team member will call interested participants to discuss the study purpose, procedures, risks, and benefits, screen for eligibility, and answer questions. Participants will be invited to our clinic to complete the consent process and begin baseline assessments.

### Research Ethics and Approval

A research assistant or project coordinator will be involved in the consenting process. The study PI will be responsible for training staff and ensuring data integrity. Research data collection will not begin until after participants provide written consent. The University of Pittsburgh Institutional Review Board approved this study’s protocol on April 19^th^, 2023 (IRB: STUDY23020169). Since this study involves minimal risk, we did not compose a data monitoring committee. This study is registered with ClinicalTrials.gov (NCT05942326).

### Randomization

Participants will be randomized at a 1:1 ratio to the Sleep GOALS intervention (n=20) or an education control group (n=20) (Figure 1). The project coordinator will generate the randomization plan using an online tool. ^49^ We will use a fixed blocked size to ensure balanced intervention groups. The randomization plan results will be stored in REDCap and only accessible by the project coordinator and a trained lifestyle coach who will support participants in the intervention arm and is not directly involved with data collection. Assignment allocation will be concealed from participants until the baseline assessments are complete. Investigators will remain blinded until post-intervention data collection is complete.

**Figure 1.**
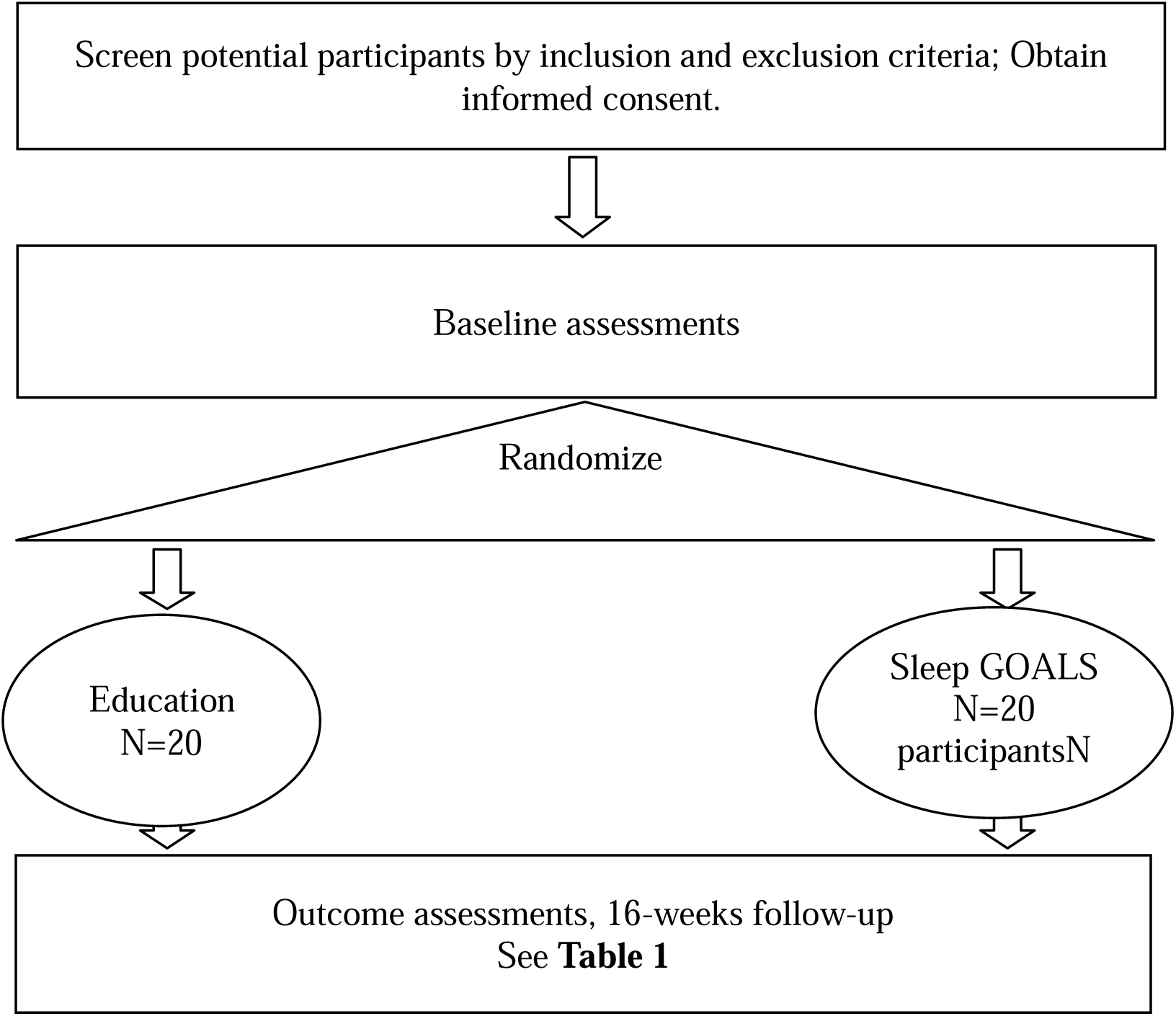
Participant flow

### Sleep GOALS

Sleep GOALS is a 16-week online intervention that includes 1) 15 to 20-minute interactive informational videos that provide strategies to improve sleep, diet, and physical activity to achieve weight loss (see Table 3), 2) a resource section, 3) a lifestyle coach, 4) a commercial activity tracker to self-monitor sleep and physical activity, and 5) a wireless scale to monitor weight. The informational videos were based on key concepts from the Transdiagnostic Sleep and Circadian Intervention^50^ and the Diabetes Prevention Program^51^ (Table 3). Participants will be able to view the material at their convenience. Each new lesson will appear five days after completing the prior one to allow participants time to process the information and implement their weekly action plan. The Sleep GOALS software will send weekly automated email alerts when a new lesson is available. The resource section includes participants’ handouts to support the action plans discussed in the videos, links to reputable resources (e.g., National Sleep Foundation, the American Heart Association), local events (e.g., 5K races), and other resources as needed for personalized support.

**Table 3.** Sleep GOALS lesson outline.

| Table 3 – Sleep GOALS lesson outline |  |
| --- | --- |
| Lesson Name | Objective |
| Week 0: Orientation | To learn how to successfully navigate the GOALS platform to access course materials. |
| Week 1: Getting Started | To learn the overall program goals and the key components of the program. |
| Week 2: Sleep Overview | To learn the processes that regulate sleep. |
| Week 3: Sleep Schedule | To learn how to create a personalized wake-up and wind-down routine |
| Week 4: Sleep Efficiency | To learn how to maximize sleep efficiency. |
| Week 5: Healthy Eating | To identify and implement USDA dietary recommendations specific to pregnant and postpartum individuals. |
| Week 6: Sleep and Eating | To learn how to improve sleep with good eating habits and vice versa. |
| Week 7: Move those Muscles | To learn the health benefits of regular physical activity. |
| Week 8: Being active as a way of life | To implement strategies for increasing daily physical activity and managing daytime dysfunction. |
| Week 9: Taking Charge of What's Around You | To learn how to modify their environment to encourage healthy eating and physical activity. |
| Week 10: Problem Solving | To learn how to apply problem-solving techniques to overcome barriers to a healthy lifestyle. |
| Week 11: Talk Back to Negative Thoughts | To learn cognitive restructuring techniques to reframe negative thoughts. |
| Week 12: Unhealthy Sleep Beliefs | To debunk common sleep myths and apply tips for better sleep. |
| Week 13: The Slippery Slope of Lifestyle Change | To learn how to identify and apply strategies to regain focus on healthy lifestyle changes after setbacks. |
| Week 14: Jump Start Your Activity Plan | To learn how to safely increase their activity intensity based on their current level. |
| Week 15: Making Social Cues Work | To learn how to identify positive social cues and integrate them into their lifestyle. |
| Week 16: Ways to Stay Motivated | To apply motivational techniques to sustain healthy lifestyle changes over the long term. |

Each participant will have a lifestyle coach who will monitor their engagement (see Table 4). They will send participants weekly messages through the intervention platform to reinforce key concepts of each lesson, respond to questions, provide personalized feedback as needed, encourage accountability, and re-engage participants who go two weeks without completing a lesson or reporting their weight, physical activity, or sleep.^52^ The coach will be a master’s level student in a health-related program (e.g., physical activity and health promotion, nutrition and dietetics, etc.). All coaches will complete a 2-day training workshop using a similar training format as the GOALS Lifestyle intervention. ^53^ To ensure treatment fidelity, the PI will randomly check 10% of each coach-participant communication through the GOALS messaging portal. The PI will meet weekly with each lifestyle coach to discuss their experience and brainstorm strategies to improve participant communication and the coach’s workflow.

**Table 4.** Primary and secondary outcomes.

| Table 4 – Primary and secondary outcomes |  |  |
| --- | --- | --- |
| Domain | Measure, Method of Assessment | Benchmarks for Success |
| Feasibility (Primary Outcome) |  |  |
| Recruitment/Enrollment | Total sample | 3-4 participants per month<br>40 total participants in 12 months |
|  | Demographics of sample | ≥30% ethnic minority |
| Retention | Attrition Rates | ≤20% loss to follow-up |
| Engagement | Modules completed | ≥14 (~80%) modules completed within 16 weeks |
|  | Weekly self-monitoring of diet, sleep, and physical activity weekly | ≥14 (~80%) weeks, ≥1 day of monitoring |
|  | Weekly weigh-ins | ≥14 (~80%) weeks, ≥1 day of weigh-ins |
|  | Total time logged in |  |
| Acceptability (Primary Outcome) |  |  |
| Acceptability | Acceptability of the intervention delivery; behavior change curricula; action plan; intervention platform | ≥80% agree or totally agree with each |
| Preliminary efficacy (Secondary Outcome) |  |  |
| Weight | Weight change | >5% weight loss |
|  | Postpartum weight retention | <5 kg of weight retention |
| The primary endpoints will inform factors related to the conduct of the study (i.e., feasibility) and the intervention content and delivery (i.e., acceptability) of the planned larger definitive efficacy trial. The secondary outcomes will provide estimates of the intervention's effects on the primary outcome of the planned larger definitive efficacy trial |  |  |

Lastly, participants will be given a wireless scale for self-monitoring weight and an activity tracker for self-monitoring sleep (daily minutes) and physical activity (daily steps, minutes). To track dietary calories, participants will be instructed to identify an app of their choice with support from the lifestyle coach. Participants will be asked to select a day and time to weigh themselves at least once weekly and enter their weight, sleep duration, daily steps, and calories into the Sleep GOALS tracking section. ^54^

### Education Control

We used the Pragmatical Model for Comparator Selection in health-related behavioral trials to guide our control group selection. ^55^ This model suggests utilizing a control group that is not likely to produce a change in the outcome of early-stage intervention development. Thus, although we considered no-treatment comparator and wait-list control conditions, we chose an education control so that participants randomized to this arm would receive a valuable, plausible condition unlikely to affect weight. Following randomization, the control condition will be emailed educational brochures from publicly available resources from the American Academy of Sleep Medicine (sleep for overall health), the National Sleep Foundation (tips for improving sleep), the American Heart Association (tips for creating a healthy eating pattern), the US Department of Agriculture Food and Nutrition Services (building a healthy eating plan while breastfeeding), and the US Department of Health and Human Services (e.g., physical activity promotion during and after pregnancy). All materials will be delivered at one time.

### Overview of Data Collection

A research assistant or project coordinator will be involved in data collection under the supervision of the PI. Data collection will include physical measurements (i.e., measured height and weight), REDCap surveys, and semi-structured interviews (Table 1). We selected previously validated interviewer-administered questionnaires relevant to the study objectives. All study data will be stored in REDCap. Hard copies, if any, will be stored in a locked filing cabinet in the PI’s university office, accessible only by authorized study team members.

#### Primary Outcomes

The primary outcomes are the intervention’s feasibility and acceptability. To systematically evaluate the feasibility and acceptability of the Sleep GOALS intervention, our study is guided by a well-established feasibility framework. This framework, adapted from the work of Bowen et al., focuses on eight domains critical to assessing health interventions: acceptability, demand, implementation, practicality, adaptation, integration, expansion, and limited-efficacy testing.^56^ By applying this multidimensional approach, we aim to thoroughly assess not only the potential for implementing Sleep GOALS on a larger scale but also identify specific areas for refinement and optimization (i.e., recruitment, retention, and engagement; Table 4). To determine acceptability, we will ask participants in the intervention arm to complete a brief 3-item REDCap survey after each lesson to rate the lesson content (e.g., was it logical, relevant, instructive) and action plan (e.g., helpful, easy to implement) on a scale of 1 to 5, where higher scores indicate higher acceptability. Lessons with an average score of <4 will be flagged for refinement.

We will then conduct 60-minute semi-structured interviews with participants at the end of the 16-week intervention. The interviews will take place in person or by video chat. We will ask participants whether the content was understandable, the action plans were achievable, and what adaptations are necessary to improve low-scoring lessons. All interviews will be audio-recorded and transcribed by trained qualitative researchers. Participants will receive $50 after completing the pre- and post-intervention assessments and a $25 honorarium after completing the semi-structured interviews.

#### Secondary Outcomes

*Weight* will be measured with a digital scale during in-person assessments with the participant wearing light clothing and no shoes at baseline and post-intervention. Postpartum weight retention will be defined as the difference in measured weight at post-intervention and self-reported pre-pregnancy weight.

#### Other Assessments

Self-reported demographic characteristics will include age, parity, breastfeeding status, race/ethnicity, marital status, number of adults in the home, level of education achieved, employment status, and annual household income. Since infant sleep/wake and feeding schedules are often cited for poor sleep quality in new mothers, we will assess these characteristics with the Infant Characteristics Questionnaire^57^ and Infant Feeding Questionnaire. ^58^ Postpartum depression will be evaluated with the Edinburgh Post Natal Depression Scale. ^59^ The Perceived Stress Scale will be used to measure perceived stress.^60^ We will use the Goals and Relative Weights Questionnaire to assess participant treatment expectations ^61^ and the Medical Outcomes Study Social Support Survey to measure social support.^62^ We will use the Barkin Index of Maternal Functioning^63^ to assess overall functioning in the past two weeks in the context of new motherhood and the American Academy of Family Physicians social determinants of health screener to identify individual needs, including housing, food, transportation, utilities, child care, employment, education, finances, and personal safety.^64^

Participants will be given a movement monitor (ActiGraph GT9X Link) to wear for seven consecutive days, 24 hours per day, at pre- and post-intervention assessments. The monitor will measure sleep and physical activity objectively. The ActiGraph GT9X Link is a triaxial accelerometer that detects vertical accelerations along three planes of movement. The total sum and patterns of movements are used to estimate physical activity and sleep. We will process the raw accelerometer data using R statistical software to assess physical activity and sleep.^65, 66^ Participants will be instructed to complete a daily sleep diary during the same monitor wear periods to help interpret the actigraphy data.^67^

*Dietary intake* will be assessed using two 24-hour recalls on random days within a seven-day window at baseline and post-intervention.^68^

### Statistical Analysis Plan

The primary objective is to assess the feasibility and acceptability of Sleep GOALS. We will assess the feasibility of the intervention by estimating rates of recruitment, retention, and engagement (i.e., weekly logins; modules completed; daily self-monitoring of diet, physical activity, and sleep; weekly monitoring of weight using point estimates (means or sample proportions) and 95% confidence intervals. We will assess acceptability by estimating satisfaction rates on various intervention components. Our benchmarks for feasibility and acceptability are similar to those used in a prior pilot trial conducted by our research team.^69^ We will use a benchmark of 80% for all feasibility and acceptability measures. To evaluate the secondary outcome, we will use descriptive statistics to compare within and between group changes in weight and postpartum weight retention. While estimating changes in sleep, diet, and physical activity is not a primary or secondary outcome, we will conduct exploratory, descriptive analysis since these are the intervention’s proposed mediators. We will use R to conduct all quantitative analyses.

An experienced qualitative research analyst will perform the qualitative analysis. Each interview will be audio-recorded and transcribed verbatim, removing personal identifiers. Then, the analyst will develop a comprehensive codebook based on the interviews. Next, two trained analysts will co-code a subset of transcripts to ensure coding reliability and establish intercoder reliability. Lastly, conventional content and thematic analyses will be applied to all transcripts, providing insights into the intervention’s acceptability. We will use Nvivo to conduct the qualitative analyses.

### Sample Size

The primary aim of the pilot RCT is to obtain estimates of feasibility and acceptability to guide the planning of a larger efficacy trial and finalize the intervention’s content and delivery. While pilot trials are not typically powered to detect statistically significant effects, enrolling 40 participants can provide sufficient data to estimate effect sizes and variability. Based on the team’s experience, a sample size of 40 (20 in each group), we can estimate 95% confidence intervals with acceptable precision (i.e., confidence interval width) of 0.31 for all feasibility and acceptability benchmarks for the entire sample and 0.44 for within-group estimates. Moreover, enrolling 40 participants balances obtaining meaningful insights and keeping within budgetary and logistical constraints.

### Participant Confidentiality

The baseline and post-intervention assessments will be conducted in a private room at our research clinic. The study participant’s contact information will be securely stored on REDCap for internal use during the study. At the end of the study, all records will remain on REDCap for as long as dictated by the reviewing IRB, Institutional policies, or sponsor requirements. Study participant research data will be stored on REDCap for purposes of statistical analysis and scientific reporting. The stored data will not include the participant’s contact or identifying information. Instead, a unique study identification number will identify individual participants and their research data. The study data entry and study management systems used by research staff will be secured and password protected.

### Adverse Event Reporting

The PI will determine whether an adverse event (AE) is expected or unexpected. An AE will be considered unexpected if the event’s nature, severity, or frequency is inconsistent with the risk information previously described for the study intervention. The occurrence of an adverse event (AE) or serious adverse event (SAE) may come to the attention of study personnel during study visits, community with the lifestyle coach during the intervention, and interviews. We will record all reportable events with start dates after informed consent is obtained until seven days (for non-serious AEs) or 30 days (for SAEs) after the last day of study participation. We will assess and document AEs and SAEs at each study visit, inquiring about the occurrence of AEs and SAEs since the previous visit and focusing on any issues related to sleep, diet, physical activity, or weight loss. We will follow up on all events for outcome information until resolution or stabilization. The PI will report AEs to the sponsor within 48 hours of the PI learning about the event.

### Protocol Deviation Reporting

The PI and research team will identify and report deviations within five working days of identification of the protocol deviation or within five working days of the scheduled protocol-required activity. All deviations will be addressed in study source documents and reported to NIH. Protocol deviations will be sent to the reviewing IRB.

### Publication Policy

This study will comply with the NIH Data Sharing Policy and Policy on the Dissemination of NIH-Funded Clinical Trial Information and the Clinical Trials Registration and Results Information Submission rule. As such, this trial is registered at ClinicalTrials.gov, and information on the results of this trial will be submitted to ClinicalTrials.gov. In addition, every attempt will be made to publish results in peer-reviewed journals. Data from this study may be requested from other researchers two years after completing the primary endpoint by contacting the PI.

## Discussion

This 16-week, single-blind pilot RCT is designed to determine the feasibility and acceptability of an internet-assisted weight loss intervention for postpartum people. Pilot testing is crucial for assessing key progression criteria to inform the feasibility and advisability of proceeding to a larger, definitive trial. These criteria encompass recruitment efficacy, participant retention, engagement levels, and preliminary changes (i.e., point estimates and measure of variability) in secondary outcomes. Our progression criteria include a set of benchmarks: successful recruitment as planned, achieving at least an 80% retention rate, high engagement with the intervention components, and indicative positive trends in secondary outcomes. These benchmarks are designed to assess the trial’s feasibility collectively, yet we acknowledge that flexibility in meeting these criteria is essential.

Our first measure of feasibility is recruitment over the planned 12-month recruitment period. We will monitor the rates and demographics of participant enrollment monthly to identify which recruitment methods yield the largest enrollment, understand potential barriers or motivators for participant enrollment, and anticipate resource allocation needs for the larger trial. If recruitment rates are below benchmarks, we will consult Clinical and Translational Science Institute (CTSI) recruitment facilitators during and after the trial to revise our recruitment protocol. When transitioning to a larger trial, these insights allow for a more streamlined and targeted recruitment process, ensuring timely participant enrollment, adequate representation, and optimal allocation of resources.

Monitoring retention rates in a pilot trial is a crucial determinant of participant engagement and the overall feasibility of the study protocol. Retention figures offer a preview of potential challenges that may arise in a full-scale trial, especially regarding participant commitment and the effectiveness of the intervention or procedure under examination. If retention rates fall below the anticipated 80% benchmark in the pilot, it signals a need to re-evaluate and adapt strategies for the larger trial. Adaptions might include enhancing participant honorarium, providing regular feedback or communication, or modifying aspects of the intervention to fit participants’ needs and preferences. By addressing these challenges head-on, based on pilot trial data, researchers can refine their approach to ensure higher retention in the subsequent larger trial, thereby enhancing the study’s validity and reliability.

We will monitor the number of modules completed, time spent logged into the Sleep GOALS platform, and behavior and weight tracking. If engagement is low, we will use semi-structured interview data to identify barriers and motivators of engagement and identify specific aspects of the intervention that may require adjustments. Likewise, we interview the lifestyle coach to discuss participant communication and obtain feedback on tools and resources, challenges, protocol fidelity, suggestions for improvement, and overall satisfaction with the program to refine the coaching protocol for the larger trial.

The secondary aim of this study is to estimate weight loss and postpartum weight retention. Exploratory aims include estimating changes in sleep, diet, and physical activity. While the study will not be sufficiently powered to detect clinically meaningful changes in weight and behavior, we will use variability estimates to inform power calculations for a larger definitive clinical trial. Likewise, the content of the intervention around physical activity and sleep will be fine-tuned based on the exploratory findings, ensuring a comprehensive, evidence-based approach that can be adaptively implemented in the subsequent trial.

### Criteria for progression to full-scale RCT

While we aim to meet all progression criteria to ensure the robustness of our pilot findings, we recognize that each criterion offers unique insights into the trial’s feasibility. Therefore, should one criterion be met—particularly recruitment—it may still warrant proceeding with a full RCT. In such instances, substantial intervention or study protocol modifications would be considered based on comprehensive feedback and data collected during the pilot phase. This approach ensures we make informed decisions on progressing to a full RCT, balancing between ideal benchmarks and practical insights gained.

### Strengths and Limitations

This study integrates critical concepts from evidenced-based sleep, diet, and physical activity interventions, potentially filling a critical research gap by informing the development of an novel weight-loss intervention for postpartum people. Moreover, the data generated from this study will allow us to capture essential data to inform recruitment, retention, data collection, and coaching protocols for a subsequent RCT. Despite the strengths of the study design, there are some notable limitations. First, while appropriate for a pilot study, the sample size limits generalizability or the ability to examine the feasibility and acceptability across sub-groups. Nonetheless, we will conduct an exploratory analysis of the association of demographic characteristics (e.g., parity, breastfeeding status, postpartum depression, race/ethnicity, relationship status, and employment status) on the intervention’s feasibility and acceptability. However, trials with larger samples will be required to provide more definitive guidance on differential response to the intervention.

In conclusion, the results of this pilot will provide significant insights into the development of definitive trials in the future. If successful, the Sleep GOALS intervention can potentially enhance weight management strategies for postpartum individuals, ultimately improving short and long-term maternal health.

## Data Availability

This is a study protocol and does not present new data

## Trial Status

The trial is currently open for recruitment, which will last through July 2024.

## Declarations

### Ethics approval and consent to participate

The University of Pittsburgh Institutional Review Board approved this study’s protocol on April 19^th^, 2023. All participants will be required to provide consent prior to enrollment and baseline assessments.

### Consent for publication

Not Applicable

### Availability of data and materials

Not Applicable

### Competing interest

Over the past three years, Dr. Daniel Buysse has been a paid or unpaid consultant to Sleep Number, Idorsia, and Eisai. All consulting agreements have been for less than $5000 per year from any single entity. Dr. Daniel Buysse is an author of the Pittsburgh Sleep Quality Index, Pittsburgh Sleep Quality Index Addendum for PTSD (PSQI-A), Brief Pittsburgh Sleep Quality Index (B-PSQI), Daytime Insomnia Symptoms Scale, Pittsburgh Sleep Diary, Insomnia Symptom Questionnaire, and RU_SATED (copyrights held by the University of Pittsburgh). These instruments have been licensed to commercial entities for fees. He is also co-author of the Consensus Sleep Diary (copyright held by Ryerson University), which is licensed to commercial entities for a fee. He has received grant support from NIH, PCORI, AHRQ, and the VA. Dr. Esa Davis is a member of the United States Preventive Services Task Force (USPSTF). Dr. Judy Chang is a member of the National Academies of Science, Engineering, and Medicine (NASEM) standing committee on Reproductive Health, Equity, and Society. This article does not necessarily represent the views and policies of the USPSTF or NASEM.

### Funding

The National Heart Lung Blood Institute 1K01 HL161439-01 funded this study.

### Authors contributions

MSH contributed to the conception and design and drafted and revised the manuscript. EMD, KZA, KMM, DJB, JCC, and MDL contributed to the manuscript’s conception and revision. MS and NK contributed to the drafting and revision of the manuscript.

## Acknowledgments

None

## Notes

### Clinical Trial

NCT05942326

### Author Declarations

The University of Pittsburgh Institutional Review Board approved this studys protocol on April 19th 2023. All participants will be required to provide consent prior to enrollment and baseline assessments.

## References

1. Vahratian A. Prevalence of overweight and obesity among women of childbearing age: results from the 2002 National Survey of Family Growth. Matern Child Health J. 2009;13(2):268–73. doi: 10.1007/s10995-008-0340-6. PubMed PMID: 18415671; PMCID: PMC2635913.

2. Flegal KM, Carroll MD, Ogden CL, Curtin LR. Prevalence and trends in obesity among US adults, 1999-2008. JAMA. 2010;303(3):235–41. doi: 10.1001/jama.2009.2014. PubMed PMID: 20071471.

3. Martin JE, Hure AJ, Macdonald-Wicks L, Smith R, Collins CE. Predictors of post-partum weight retention in a prospective longitudinal study. Matern Child Nutr. 2014;10(4):496–509. Epub 2012/09/15. doi: 10.1111/j.1740-8709.2012.00437.x. PubMed PMID: 22974518.

4. Bogaerts A, De Baetselier E, Ameye L, Dilles T, Van Rompaey B, Devlieger R. Postpartum weight trajectories in overweight and lean women. Midwifery. 2017;49:134–41. Epub 2016/09/18. doi: 10.1016/j.midw.2016.08.010. PubMed PMID: 27638342.

5. Rooney BL, Schauberger CW. Excess pregnancy weight gain and long-term obesity: one decade later. Obstet Gynecol. 2002;100(2):245–52. PubMed PMID: 12151145.

6. Rooney BL, Schauberger CW, Mathiason MA. Impact of perinatal weight change on long-term obesity and obesity-related illnesses. Obstet Gynecol. 2005;106(6):1349–56. Epub 2005/12/02. doi: 10.1097/01.AOG.0000185480.09068.4a. PubMed PMID: 16319262.

7. Bartsch E, Medcalf KE, Park AL, Ray JG, High Risk of Pre-eclampsia Identification G. Clinical risk factors for pre-eclampsia determined in early pregnancy: systematic review and meta-analysis of large cohort studies. BMJ. 2016;353:i1753. Epub 2016/04/21. doi: 10.1136/bmj.i1753. PubMed PMID: 27094586; PMCID: PMC4837230.

8. Torloni MR, Betran AP, Horta BL, Nakamura MU, Atallah AN, Moron AF, Valente O. Prepregnancy BMI and the risk of gestational diabetes: a systematic review of the literature with meta-analysis. Obes Rev. 2009;10(2):194–203. Epub 2008/12/06. doi: 10.1111/j.1467-789X.2008.00541.x. PubMed PMID: 19055539.

9. Force USPST, Curry SJ, Krist AH, Owens DK, Barry MJ, Caughey AB, Davidson KW, Doubeni CA, Epling JW, Jr., Grossman DC, Kemper AR, Kubik M, Landefeld CS, Mangione CM, Phipps MG, Silverstein M, Simon MA, Tseng CW, Wong JB. Behavioral Weight Loss Interventions to Prevent Obesity-Related Morbidity and Mortality in Adults: US Preventive Services Task Force Recommendation Statement. JAMA. 2018;320(11):1163–71. Epub 2018/10/17. doi: 10.1001/jama.2018.13022. PubMed PMID: 30326502.

10. Nascimento SL, Pudwell J, Surita FG, Adamo KB, Smith GN. The effect of physical exercise strategies on weight loss in postpartum women: a systematic review and meta-analysis. Int J Obes (Lond). 2014;38(5):626–35. doi: 10.1038/ijo.2013.183. PubMed PMID: 24048142.

11. Lim S, O’Reilly S, Behrens H, Skinner T, Ellis I, Dunbar JA. Effective strategies for weight loss in post-partum women: a systematic review and meta-analysis. Obes Rev. 2015;16(11):972–87. Epub 2015/08/28. doi: 10.1111/obr.12312. PubMed PMID: 26313354.

12. Ferrara A, Hedderson MM, Brown SD, Albright CL, Ehrlich SF, Tsai AL, Caan BJ, Sternfeld B, Gordon NP, Schmittdiel JA, Gunderson EP, Mevi AA, Herman WH, Ching J, Crites Y, Quesenberry CP, Jr. The Comparative Effectiveness of Diabetes Prevention Strategies to Reduce Postpartum Weight Retention in Women With Gestational Diabetes Mellitus: The Gestational Diabetes’ Effects on Moms (GEM) Cluster Randomized Controlled Trial. Diabetes Care. 2016;39(1):65–74. Epub 2015/12/15. doi: 10.2337/dc15-1254. PubMed PMID: 26657945; PMCID: PMC4686847.

13. Herring SJ, Cruice JF, Bennett GG, Darden N, Wallen JJ, Rose MZ, Davey A, Foster GD. Intervening during and after pregnancy to prevent weight retention among African American women. Prev Med Rep. 2017;7:119–23. Epub 2017/07/01. doi: 10.1016/j.pmedr.2017.05.015. PubMed PMID: 28660118; PMCID: PMC5479961.

14. Holmes VA, Draffin CR, Patterson CC, Francis L, Irwin J, McConnell M, Farrell B, Brennan SF, McSorley O, Wotherspoon AC, Davies M, McCance DR, Group PS. Postnatal Lifestyle Intervention for Overweight Women With Previous Gestational Diabetes: A Randomized Controlled Trial. J Clin Endocrinol Metab. 2018;103(7):2478–87. Epub 2018/05/16. doi: 10.1210/jc.2017-02654. PubMed PMID: 29762737.

15. Craigie AM, Macleod M, Barton KL, Treweek S, Anderson AS, WeighWell t. Supporting postpartum weight loss in women living in deprived communities: design implications for a randomised control trial. Eur J Clin Nutr. 2011;65(8):952–8. Epub 2011/05/12. doi: 10.1038/ejcn.2011.56. PubMed PMID: 21559034; PMCID: PMC3154650.

16. National Sleep Foundation. Women and Sleep: Summary Findings of 2007 Sleep in America Poll 2007 [cited 2016 May 13]. Available from: https://sleepfoundation.org/sleep-polls-data/sleep-in-america-poll/2007-women-and-sleep.

17. Kang MJ, Matsumoto K, Shinkoda H, Mishima M, Seo YJ. Longitudinal study for sleep-wake behaviours of mothers from pre-partum to post-partum using actigraph and sleep logs. Psychiatry Clin Neurosci. 2002;56(3):251–2. doi: 10.1046/j.1440-1819.2002.00992.x. PubMed PMID: 12047581.

18. Lee KA, Zaffke ME, McEnany G. Parity and sleep patterns during and after pregnancy. Obstet Gynecol. 2000;95(1):14–8. PubMed PMID: 10636494.

19. Gunderson EP, Rifas-Shiman SL, Oken E, Rich-Edwards JW, Kleinman KP, Taveras EM, Gillman MW. Association of fewer hours of sleep at 6 months postpartum with substantial weight retention at 1 year postpartum. Am J Epidemiol. 2008;167(2):178–87. doi: 10.1093/aje/kwm298. PubMed PMID: 17971337; PMCID: PMC2930882.

20. Taveras EM, Rifas-Shiman SL, Rich-Edwards JW, Gunderson EP, Stuebe AM, Mantzoros CS. Association of maternal short sleep duration with adiposity and cardiometabolic status at 3 years postpartum. Obesity (Silver Spring). 2011;19(1):171–8. doi: 10.1038/oby.2010.117. PubMed PMID: 20489690; PMCID: PMC3099421.

21. Althuizen E, van Poppel MN, de Vries JH, Seidell JC, van Mechelen W. Postpartum behaviour as predictor of weight change from before pregnancy to one year postpartum. BMC Public Health. 2011;11:165. Epub 2011/03/18. doi: 10.1186/1471-2458-11-165. PubMed PMID: 21410953; PMCID: PMC3068095.

22. Siega-Riz AM, Herring AH, Carrier K, Evenson KR, Dole N, Deierlein A. Sociodemographic, perinatal, behavioral, and psychosocial predictors of weight retention at 3 and 12 months postpartum. Obesity (Silver Spring). 2010;18(10):1996–2003. doi: 10.1038/oby.2009.458. PubMed PMID: 20035283; PMCID: PMC2902688.

23. Strand LB, Laugsand LE, Wisloff U, Nes BM, Vatten L, Janszky I. Insomnia symptoms and cardiorespiratory fitness in healthy individuals: the Nord-Trondelag Health Study (HUNT). Sleep. 2013;36(1):99–108. Epub 2013/01/05. doi: 10.5665/sleep.2310. PubMed PMID: 23288976; PMCID: PMC3524509.

24. Chasens ER, Sereika SM, Weaver TE, Umlauf MG. Daytime sleepiness, exercise, and physical function in older adults. J Sleep Res. 2007;16(1):60–5. Epub 2007/02/21. doi: 10.1111/j.1365-2869.2007.00576.x. PubMed PMID: 17309764.

25. Holfeld B, Ruthig JC. A longitudinal examination of sleep quality and physical activity in older adults. J Appl Gerontol. 2014;33(7):791–807. Epub 2014/09/19. doi: 10.1177/0733464812455097. PubMed PMID: 25231754.

26. Haario P, Rahkonen O, Laaksonen M, Lahelma E, Lallukka T. Bidirectional associations between insomnia symptoms and unhealthy behaviours. J Sleep Res. 2013;22(1):89–95. Epub 2012/09/18. doi: 10.1111/j.1365-2869.2012.01043.x. PubMed PMID: 22978579.

27. Lambiase MJ, Gabriel KP, Kuller LH, Matthews KA. Temporal relationships between physical activity and sleep in older women. Med Sci Sports Exerc. 2013;45(12):2362–8. Epub 2013/06/07. doi: 10.1249/MSS.0b013e31829e4cea. PubMed PMID: 23739529; PMCID: PMC3833970.

28. Baron KG, Reid KJ, Kern AS, Zee PC. Role of sleep timing in caloric intake and BMI. Obesity (Silver Spring). 2011;19(7):1374–81. Epub 2011/04/30. doi: 10.1038/oby.2011.100. PubMed PMID: 21527892.

29. Garaulet M, Gomez-Abellan P, Alburquerque-Bejar JJ, Lee YC, Ordovas JM, Scheer FA. Timing of food intake predicts weight loss effectiveness. Int J Obes (Lond). 2013;37(4):604–11. Epub 2013/01/30. doi: 10.1038/ijo.2012.229. PubMed PMID: 23357955; PMCID: PMC3756673.

30. McHill AW, Phillips AJ, Czeisler CA, Keating L, Yee K, Barger LK, Garaulet M, Scheer FA, Klerman EB. Later circadian timing of food intake is associated with increased body fat. Am J Clin Nutr. 2017;106(5):1213–9. Epub 2017/09/08. doi: 10.3945/ajcn.117.161588. PubMed PMID: 28877894; PMCID: PMC5657289.

31. Wang JB, Patterson RE, Ang A, Emond JA, Shetty N, Arab L. Timing of energy intake during the day is associated with the risk of obesity in adults. J Hum Nutr Diet. 2014;27 Suppl 2:255–62. Epub 2013/07/03. doi: 10.1111/jhn.12141. PubMed PMID: 23808897.

32. Jakubowicz D, Barnea M, Wainstein J, Froy O. High caloric intake at breakfast vs. dinner differentially influences weight loss of overweight and obese women. Obesity (Silver Spring). 2013;21(12):2504–12. Epub 2013/03/21. doi: 10.1002/oby.20460. PubMed PMID: 23512957.

33. Chaix A, Manoogian ENC, Melkani GC, Panda S. Time-Restricted Eating to Prevent and Manage Chronic Metabolic Diseases. Annu Rev Nutr. 2019;39:291–315. Epub 2019/06/11. doi: 10.1146/annurev-nutr-082018-124320. PubMed PMID: 31180809; PMCID: PMC6703924.

34. Rihm JS, Menz MM, Schultz H, Bruder L, Schilbach L, Schmid SM, Peters J. Sleep Deprivation Selectively Upregulates an Amygdala-Hypothalamic Circuit Involved in Food Reward. J Neurosci. 2019;39(5):888–99. Epub 2018/12/19. doi: 10.1523/JNEUROSCI.0250-18.2018. PubMed PMID: 30559151; PMCID: PMC6382977.

35. Fang Z, Spaeth AM, Ma N, Zhu S, Hu S, Goel N, Detre JA, Dinges DF, Rao H. Altered salience network connectivity predicts macronutrient intake after sleep deprivation. Sci Rep. 2015;5:8215. Epub 2015/02/04. doi: 10.1038/srep08215. PubMed PMID: 25645575; PMCID: PMC4314629.

36. Yang CL, Schnepp J, Tucker RM. Increased Hunger, Food Cravings, Food Reward, and Portion Size Selection after Sleep Curtailment in Women Without Obesity. Nutrients. 2019;11(3). Epub 2019/03/22. doi: 10.3390/nu11030663. PubMed PMID: 30893841; PMCID: PMC6470707.

37. Ness KM, Strayer SM, Nahmod NG, Schade MM, Chang AM, Shearer GC, Buxton OM. Four nights of sleep restriction suppress the postprandial lipemic response and decrease satiety. J Lipid Res. 2019. Epub 2019/09/06. doi: 10.1194/jlr.P094375. PubMed PMID: 31484696.

38. Logue EE, Bourguet CC, Palmieri PA, Scott ED, Matthews BA, Dudley P, Chipman KJ. The better weight-better sleep study: a pilot intervention in primary care. Am J Health Behav. 2012;36(3):319–34. Epub 2012/03/01. doi: 10.5993/AJHB.36.3.4. PubMed PMID: 22370434.

39. Diabetes Prevention Program Research G. The Diabetes Prevention Program (DPP): description of lifestyle intervention. Diabetes Care. 2002;25(12):2165–71. Epub 2002/11/28. doi: 10.2337/diacare.25.12.2165. PubMed PMID: 12453955; PMCID: PMC1282458.

40. Henderson JM, France KG, Owens JL, Blampied NM. Sleeping through the night: the consolidation of self-regulated sleep across the first year of life. Pediatrics. 2010;126(5):e1081–7. Epub 2010/10/27. doi: 10.1542/peds.2010-0976. PubMed PMID: 20974775.

41. American College of Obstetricians and Gynecologists. Exercise during pregnancy and the postpartum period. Obstetrics and gynecology. 2002;99:171–3.

42. Warburton D, Jamnik V, Bredin S, Gledhill N. The Physical Activity Readiness Questionnaire for Everyone (PAR-Q+) and Electronic Physical Activity Readiness Medical Examination (ePARmed-X+). The Health & Fitness Journal of Canada 2011;4(2):3–17. doi: 10.14288/hfjc.v4i2.103.

43. Ravyts SG, Dzierzewski JM, Perez E, Donovan EK, Dautovich ND. Sleep Health as Measured by RU SATED: A Psychometric Evaluation. Behav Sleep Med. 2021;19(1):48–56. Epub 2019/12/13. doi: 10.1080/15402002.2019.1701474. PubMed PMID: 31829724; PMCID: PMC7289662.

44. Siddiqui A, Azria E, Egorova N, Deneux-Tharaux C, Howell EA. Contribution of Prepregnancy Obesity to Racial and Ethnic Disparities in Severe Maternal Morbidity. Obstetrics and gynecology. 2021;137(5):864–72. doi: 10.1097/AOG.0000000000004356. PubMed PMID: 33831920; PMCID: PMC9725890.

45. Headen IE, Davis EM, Mujahid MS, Abrams B. Racial-ethnic differences in pregnancy-related weight. Adv Nutr. 2012;3(1):83–94. Epub 20120105. doi: 10.3945/an.111.000984. PubMed PMID: 22332106; PMCID: PMC3262620.

46. Pitt Clinical and Translational Science Institute. Pitt+Me [cited 2023 2023]. Available from: https://pittplusme.org/.

47. Magee-Womens Research Institute. Magee Obstetric Maternal & Infant (MOMI) Database and Biobank, 2024 [cited 2024 2024]. Available from: https://mageewomens.org/for-researchers/core-facilities/magee-obstetric-maternal-infant-momi-database-and-biobank?utm_mrid=mrid3773&utm_source=GOOGLE&utm_medium=cpc&utm_campaign=71700000107185291&utm_adgroup=58700008292633458&utm_term=pregnancy+biobank&utm_advertiserid=700000002768137&gclid=Cj0KCQjwn7mwBhCiARIsAGoxjaKHWL_oaS6PUoQf5LeX_57ecZY0R6ZHLW4gHWA4R-RzcbdVt4lb6MoaAmLnEALw_wcB&gclsrc=aw.ds.

48. Pitt Clinical and Translational Science Institute. Newborn Research Support Service 2024 [cited 2024 April]. Available from: https://ctsi.pitt.edu/research-services/research-facilities-networks/newborn-research-support-service/.

49. Urbaniak G, S P. Research Randomizer (Version 4.0) [Computer Software] 2013 [cited 2016 Feb]. Available from: www.randomizer.org.

50. Harvey AG. A transdiagnostic approach to treating sleep disturbance in psychiatric disorders. Cogn Behav Ther. 2009;38 Suppl 1:35–42. doi: 10.1080/16506070903033825. PubMed PMID: 19697179.

51. McTigue KM, Conroy MB, Hess R, Bryce CL, Fiorillo AB, Fischer GS, Milas NC, Simkin-Silverman LR. Using the internet to translate an evidence-based lifestyle intervention into practice. Telemed J E Health. 2009;15(9):851–8. Epub 2009/11/19. doi: 10.1089/tmj.2009.0036. PubMed PMID: 19919191.

52. Lyden JR, Zickmund SL, Bhargava TD, Bryce CL, Conroy MB, Fischer GS, Hess R, Simkin-Silverman LR, McTigue KM. Implementing health information technology in a patient-centered manner: patient experiences with an online evidence-based lifestyle intervention. J Healthc Qual. 2013;35(5):47–57. Epub 2013/09/06. doi: 10.1111/jhq.12026. PubMed PMID: 24004039.

53. Simkin-Silverman LR, Conroy MB, Bhargava T, McTigue KM. Development of an online diabetes prevention lifestyle intervention coaching protocol for use in primary care practice. Diabetes Educ. 2011;37(2):263–8. Epub 2011/03/23. doi: 10.1177/0145721710396587. PubMed PMID: 21421991.

54. Madigan CD, Daley AJ, Lewis AL, Aveyard P, Jolly K. Is self-weighing an effective tool for weight loss: a systematic literature review and meta-analysis. Int J Behav Nutr Phys Act. 2015;12:104. Epub 2015/08/22. doi: 10.1186/s12966-015-0267-4. PubMed PMID: 26293454; PMCID: PMC4546162.

55. Freedland KE, King AC, Ambrosius WT, Mayo-Wilson E, Mohr DC, Czajkowski SM, Thabane L, Collins LM, Rebok GW, Treweek SP, Cook TD, Edinger JD, Stoney CM, Campo RA, Young-Hyman D, Riley WT, National Institutes of Health Office of B, Social Sciences Research Expert Panel on Comparator Selection in B, Social Science Clinical T. The selection of comparators for randomized controlled trials of health-related behavioral interventions: recommendations of an NIH expert panel. J Clin Epidemiol. 2019;110:74–81. Epub 2019/03/04. doi: 10.1016/j.jclinepi.2019.02.011. PubMed PMID: 30826377; PMCID: PMC6543841.

56. Bowen DJ, Kreuter M, Spring B, Cofta-Woerpel L, Linnan L, Weiner D, Bakken S, Kaplan CP, Squiers L, Fabrizio C, Fernandez M. How we design feasibility studies. Am J Prev Med. 2009;36(5):452–7. doi: 10.1016/j.amepre.2009.02.002. PubMed PMID: 19362699; PMCID: PMC2859314.

57. Kempler L, Sharpe L, Bartlett D. Sleep education during pregnancy for new mothers. BMC Pregnancy Childbirth. 2012;12:155. doi: 10.1186/1471-2393-12-155. PubMed PMID: 23244163; PMCID: PMC3546917.

58. Lakshman RR, Landsbaugh JR, Schiff A, Hardeman W, Ong KK, Griffin SJ. Development of a questionnaire to assess maternal attitudes towards infant growth and milk feeding practices. Int J Behav Nutr Phys Act. 2011;8:35. doi: 10.1186/1479-5868-8-35. PubMed PMID: 21510900; PMCID: PMC3111341.

59. Cox JL, Holden JM, Sagovsky R. Detection of postnatal depression. Development of the 10-item Edinburgh Postnatal Depression Scale. The British journal of psychiatry : the journal of mental science. 1987;150:782–6. PubMed PMID: 3651732.

60. Cohen S, Kamarck T, Mermelstein R. A global measure of perceived stress. J Health Soc Behav. 1983;24(4):385–96. PubMed PMID: 6668417.

61. Foster GD, Wadden TA, Vogt RA, Brewer G. What is a reasonable weight loss? Patients’ expectations and evaluations of obesity treatment outcomes. J Consult Clin Psychol. 1997;65(1):79–85. Epub 1997/02/01. doi: 10.1037//0022-006x.65.1.79. PubMed PMID: 9103737.

62. Sherbourne CD, Stewart AL. The MOS social support survey. Soc Sci Med. 1991;32(6):705–14. Epub 1991/01/01. doi: 10.1016/0277-9536(91)90150-b. PubMed PMID: 2035047.

63. Barkin JL, Wisner KL, Bromberger JT, Beach SR, Terry MA, Wisniewski SR. Development of the Barkin Index of Maternal Functioning. J Womens Health (Larchmt). 2010;19(12):2239–46. Epub 20101105. doi: 10.1089/jwh.2009.1893. PubMed PMID: 21054183; PMCID: PMC3003914.

64. American Academy of Family Physicians. Social needs screening tool 2018 [cited 2024 2024]. Available from: https://www.aafp.org/family-physician/patient-care/the-everyone-project/toolkit.html.

65. Hart TL, Swartz AM, Cashin SE, Strath SJ. How many days of monitoring predict physical activity and sedentary behaviour in older adults? Int J Behav Nutr Phys Act. 2011;8:62. Epub 2011/06/18. doi: 10.1186/1479-5868-8-62. PubMed PMID: 21679426; PMCID: 3130631.

66. Matthews CE, Ainsworth BE, Thompson RW, Bassett DR, Jr. Sources of variance in daily physical activity levels as measured by an accelerometer. Medicine and science in sports and exercise. 2002;34(8):1376–81.

67. Carney CE, Buysse DJ, Ancoli-Israel S, Edinger JD, Krystal AD, Lichstein KL, Morin CM. The consensus sleep diary: standardizing prospective sleep self-monitoring. Sleep. 2012;35(2):287–302. Epub 2012/02/02. doi: 10.5665/sleep.1642. PubMed PMID: 22294820; PMCID: PMC3250369.

68. Subar AF, Kirkpatrick SI, Mittl B, Zimmerman TP, Thompson FE, Bingley C, Willis G, Islam NG, Baranowski T, McNutt S, Potischman N. The Automated Self-Administered 24-hour dietary recall (ASA24): a resource for researchers, clinicians, and educators from the National Cancer Institute. J Acad Nutr Diet. 2012;112(8):1134–7. doi: 10.1016/j.jand.2012.04.016. PubMed PMID: 22704899; PMCID: PMC3721511.

69. Sherifali D, Hess R, McTigue KM, Brozic A, Ng K, Gerstein H. Evaluating the feasibility and impact of an internet-based lifestyle management program in a diabetes care setting. Diabetes Technol Ther. 2014;16(6):358–62. Epub 2014/03/04. doi: 10.1089/dia.2013.0278. PubMed PMID: 24580377.

